# its2s: a Python package for two-stage interrupted time series analysis using machine learning

**DOI:** 10.64898/2026.07.02.26357175

**Authors:** Lauren Blair Wilner, Joan A. Casey, Stephen J. Mooney, Vivian Do, Yiqun Ma, Tarik Benmarhnia, Arnab K. Dey

**Affiliations:** Department of Epidemiology, University of Washington School of Public Health, Seattle, WA, USA; Department of Environmental and Occupational Health Sciences, University of Washington School of Public Health, Seattle, WA, USA; Department of Environmental Health Sciences, Columbia University Mailman School of Public Health, New York, NY, USA; Huntsman Cancer Institute, University of Utah, Salt Lake City, UT, USA; Scripps Institution of Oceanography, University of California San Diego, La Jolla, CA, USA; Irset Institut de Recherche en Santé, Environnement et Travail, UMR-S 1085, Inserm, University of Rennes, EHESP, Rennes, France

**Author notes:** **Corresponding Author**: Lauren Blair Wilner, MPH, Department of Epidemiology, University of Washington School of Public Health Seattle, WA 98195. **Data availability**: All code and data are available in the GitHub repository.

## Abstract

When randomized controlled trials are infeasible, researchers may leverage natural experiments for causal inference. Interrupted time-series (ITS) designs compare observed post-event trends to counterfactual predictions from pre-event data. Two-stage ITS designs use flexible models to generate optimized counterfactual predictions in the first stage, then estimate intervention effects by comparing observed to predicted outcomes in the second stage. Fitting high-dimensional versions of these models is challenging, requiring systematic infrastructure to ensure rigor and reproducibility. In response, we developed `its2s`, an open-source Python package implementing the two-stage ITS design with machine learning. `its2s` allows users to specify an intervention date and training/testing periods, select among built-in model architectures (e.g., Prophet-XGBoost, NeuralProphet), and generate confidence intervals via moving block bootstrap, preserving temporal autocorrelation in residuals. `its2s` layers defaults, configuration files, and runtime overrides to support workflows ranging from rapid default implementations to highly tailored analyses. We validated `its2s` using two case studies: a simulation with a 12% policy effect, recovering the true effect as 11.77%, and an analysis of the 2021 Pacific Northwest heat dome, finding 53% excess injury mortality over the following three weeks. `its2s` provides a flexible, reproducible framework for ITS-based quasi-experimental research, lowering barriers to rigorous machine learning-based counterfactual modeling.

## 2 Introduction

Causal inference is a central goal in health and policy research, where the causal effect is defined as the contrast between outcomes under treatment and outcomes under no treatment for the same units. ^1,2^ A randomized controlled trial (RCT) is often considered the gold standard for internal validity, because well-conducted randomization creates two exchangeable groups and researchers control treatment and assessment, allowing them to infer that differences in the outcome can be attributed to the treatment. However, in multiple settings, including environmental exposures or social policies, RCTs are often infeasible or unethical. ^3,4^ For example, researchers cannot assign climate disaster exposure to one group and a placebo to another – ruling out an RCT and making causal effects difficult to identify. In these cases, researchers may leverage natural experiments – events such as policy interventions, climate disasters, or disease outbreaks – whose timing is plausibly exogenous to factors driving the trends of a given outcome of interest, thus approximating randomization. ^4^ Some quasi-experimental methods exploit this exogeneity to estimate causal effects from observational data, with different designs relying on different strategies to reconstruct the missing counterfactual. Interrupted time series (ITS) designs do so by modeling the outcome’s pre-event trend and attributing post-event deviations from that trend to the event of interest. ITS designs are particularly valuable when no credibly comparable control units are available – as is common for climate disasters such as wildfires and heatwaves, and for broad-coverage public health policies. ^3,5,6^

ITS designs are applied to time series data: repeated measurements of an aggregate outcome -– such as weekly deaths or daily hospitalizations – collected at regular intervals in a population before and after an event. Beyond random variation, such series typically exhibit systematic temporal structure, including secular trends, seasonality, and temporal autocorrelation. Fundamentally, ITS designs compare observed post-event outcomes to counterfactual post-event outcomes – what would have been expected in the absence of the event. ^6^ To estimate this counterfactual, the pre-event portion of the series is used to characterize the temporal structure, which is then projected into the post-event period. The most common implementation uses segmented regression to jointly model pre- and post-event trends as two separate linear segments. ^5,7^ This generates estimates of both immediate level shifts (intercept changes) and longer-term trend shifts (slope changes) following the event. ^6^

Besides traditional identification assumptions in causal inference (e.g. correct model specification, no spillover), using ITS designs requires additional assumptions including (1) the event timing is clearly defined, and (2) no other major events sharply affecting the outcome occur concurrently with the event of interest. ^6^ Additionally, assumptions about the form of the counterfactual depend on the specific ITS design. In single-group ITS designs (sometimes called single ITS, or SITS), where no contemporaneous control series is available, the design assumes that when projected forward, the pre-event trend accurately represents what would have occurred in the absence of the event. Empirical work has shown that violations of this within-unit counterfactual assumption can be consequential. ^8^

Beyond these design-level identification assumptions, the most common ITS implementation, segmented regression, has model-specific limitations. First, the researcher must pre-specify the functional form of the underlying trend (typically linear, with optional seasonal or polynomial terms) and the intervention effect (e.g., an abrupt level shift, a gradual slope change, or a lagged response) by hard-coding the corresponding dummy and interaction terms into the model. ^3,5,9^ Selecting the functional form after inspecting the post-event data increases the risk of spurious conclusions and has been identified as a primary threat to ITS validity. ^6,8^Second, positively autocorrelated residuals – common in epidemiological time series – deflate standard errors and produce anti-conservative confidence intervals. While corrections such as Prais-Winsten or Restricted Maximum Likelihood with a Satterthwaite degrees-of-freedom adjustment can restore near-nominal coverage in longer series, no method reliably does so in the short series typical of applied ITS studies. ^10,11^ Third, segmented regression returns a small set of global parameters (level and slope changes) rather than time-unit-specific excess estimates, limiting its utility for high-dimensional or pooled analyses. To address these limitations, several alternatives have been proposed, including Autoregressive Integrated Moving Average (ARIMA/ARIMAX), Bayesian ITS, and two-stage approaches with machine learning (ML). ^7,9,12^

The two-stage ITS approach with ML is a flexible alternative to single-group ITS models that addresses the limitations specific to segmented regression. ^9,13^The “two-stage” label refers to a deliberate separation of tasks. In the first stage, a flexible ML model (e.g., a hybrid Prophet-XGBoost model) is trained on a subset of pre-event data, with the remaining pre-event data held out for testing and validation. Once a model is selected based on predictive performance, it is used to forecast the post-event period under the counterfactual scenario that pre-event trends would continue absent the event. In the second stage, researchers calculate the effect of the event by comparing the observed post-event series to this counterfactual. Separating these stages strengthens the analysis because (1) since the first-stage model never sees post-event data, the functional form cannot be tuned to the effect it is meant to estimate, and (2) the comparison is made at every time point, yielding a full trajectory of excess estimates rather than one or two global coefficients. Using an ML model further strengthens the credibility of the counterfactual, as flexible learners handle nonlinear trends, seasonality, and autocorrelation without pre-specification of the functional form. As with all single-group designs, the two-stage ML approach inherits the within-unit counterfactual assumption discussed above. However, it avoids the additional assumption required by controlled ITS designs that exposed and unexposed groups have parallel outcome trajectories over time. This is a meaningful advantage when no credible control unit exists. ML flexibility strengthens the credibility of the within-unit extrapolation, but it does not eliminate the assumption. Equally important, because model selection in stage 1 relies only on pre-event data, the impact model is never trained on (and thus never sees) post-event outcomes – structurally removing the data-driven specification bias that can compromise traditional ITS. The two-stage approach also yields time-unit-specific excess estimates rather than a single level or slope coefficient, allowing for richer characterization of dynamic and cumulative effects. ^13^

Technical and computational barriers may limit uptake of the two-stage ITS approach with ML. Most applied researchers are experts in their respective fields but have limited specialized training in selecting cross-validation strategies, splitting data into training and testing sets, or choosing appropriate ML architectures for time series forecasting. ML approaches are also computationally demanding, and their complexity can make the underlying modeling assumptions difficult to state and test. ^9^ Generating valid confidence intervals adds further complexity – methods such as moving block bootstrap (MBB) preserve temporal dependence by resampling contiguous blocks of residuals rather than individual observations, but require careful selection of block length, which is rarely straightforward. ^9,13^ When researchers seek to evaluate multiple events, outcomes, or model specifications, tracking versions and ensuring reproducibility becomes challenging. While analytic code accompanying published applications of the two-stage ML ITS approach is publicly available, ^13^ to our knowledge, no general-purpose, open-source software package implementing this design exists. Such a package could broaden adoption of the method and more reproducible research.

We developed `its2s`, an open-source Python package that provides a user-friendly implementation of the two-stage ITS approach with ML. The package couples built-in forecasting architectures with fully customizable default configurations. Reproducibility is supported through timestamped, versioned run directories and deterministic seeding across parallel batch runs. Here, we describe the methodological foundation for the two-stage ITS with ML design; detail the `its2s` package implementing this approach, and demonstrate its application through two case studies.

## 3 Methods

To make the two-stage ITS with ML approach accessible to researchers, we developed `its2s`, an open-source Python package that implements this design with minimal barriers to entry. Users can provide as little as a time series dataset and specification of intervention/event date, leaning on built-in defaults and automation of model fitting, comparison, and effect estimation in `its2s`. Researchers can also leverage the full flexibility of `its2s` to customize every aspect of the workflow from data splitting and cross-validation strategies to hyperparameter tuning ranges and model selection. This flexibility allows researchers to conduct analyses ranging from rapid default implementations to fully customized workflows tailored to their specific research questions, all within a reproducible framework.

In stage 1, `its2s` splits the portion of the time series preceding the event into a training window and a held-out testing window. A ML model is trained on the training window to learn the outcome’s temporal patterns, then evaluated on the testing window to assess out-of-sample predictive power. Since the goal of this model is to forecast the unobservable counterfactual post-event trend, for which we do not know the truth, this evaluation of out-of-sample predictive power is essential. Flexible models can fit their training data well while generalizing poorly, ^14–16^ so it is therefore vital to ensure that the counterfactual accurately represents the expected post-event period had the event not occurred. Train/test separation of this kind is the standard approach for evaluating predictive models in ML and time series forecasting; `its2s` makes it a structural, non-optional step of the ITS workflow. ^16–19^

The selected model is then used to generate counterfactual predictions for the post-event period (or the holdout, in ML terms) - the expected time series had the event not occurred. In stage 2, `its2s` compares observed post-event outcomes to these counterfactual predictions to estimate the effect attributable to the event. Uncertainty is quantified using moving block bootstraps (MBB), ^20^ which resamples blocks of residuals rather than individual observations to preserve temporal autocorrelation, following the approach in Dey et al. ^13^ The statistical theory underlying the two-stage ITS approach is described in more detail in Dey et al. ^13^

The two stages of the ITS are implemented in `its2s` through a small set of high-level functions organized around a simple core workflow. The primary entry point is `run_single_its()`, which executes the full pipeline for a single outcome series. This function accepts a time series dataframe, an event date, and optional specifications for the target, date, and covariate columns, the forecasting model, a YAML config (via path or runtime overrides), an output directory, and a random seed. The function then splits the pre-event period into training and test windows and fits and evaluates the selected forecasting model in Stage 1. Currently, this package supports the following ML model architectures for this model: (1) Prophet + XG-Boost hybrid, (2) NeuralProphet, (3) Prophet then XGBoost, (4) ARIMA. As part of the Stage 1 fit, `its2s` automatically runs residual diagnostic checks via `compute_diagnostics()`, which evaluates the Ljung-Box statistic at pre-defined lags to detect residual autocorrelation ^17^ and applies the Shapiro-Wilk test for normality. ^21^ These diagnostics directly assess the validity of the moving-block bootstrap used in Stage 2, whose block length governs how much residual autocorrelation the resampling preserves and therefore the coverage of the resulting intervals. The block length may be fixed by the user (the default, specified in observations), or estimated automatically using the Politis and White plug-in rule ^22^; in addition, `its2s` provides a diagnostic that traces the width of the event-window confidence interval across a range of candidate block lengths, letting the user confirm that the selected length falls in a region where interval width has stabilized. Because no block length restores nominal coverage when the Stage 1 residuals remain strongly autocorrelated, the Ljung-Box diagnostic serves to verify that the fitted model has adequately captured the temporal dependence before the bootstrap is applied. The pipeline returns a `PipelineResult` object containing fitted values, effect estimates, bootstrap confidence intervals, and model diagnostics. All results are accompanied by `plot_counterfactual()`, which produces a standardized figure showing the observed series, in-sample model fit, counterfactual prediction with confidence interval ribbon, and event marker. In the case where the same outcome is studied across multiple geographic units or sub-groups, users can use `run_batch()` to produce a list of series, parallelizing execution across cores and writing outputs to automatically versioned run directories that encode the run date and bootstrap simulation count for all combinations. Configuration at every step is managed through a YAML file loaded via `load_config()`, which specifies the split method, bootstrap parameters, and model hyperparameters; any parameter can be overridden at runtime via the `config_overrides` argument, and all overrides are documented in the model outputs, providing a reproducible specification that can be version-controlled alongside the data and code.

For researchers who want to interrogate model selection before committing to a final run, `its2s` also provides a set of pre-analysis tools: `compare_models()` runs all available model architectures on the same series and returns a ranked comparison table, `tune_model()` searches the hyperparameter space for a chosen model via Latin hypercube sampling, and `time_series_cv()` evaluates a single model’s out-of-sample performance across expanding folds independently of a full pipeline run.

We illustrate the package’s application in two case studies. First, we evaluate `its2s` using a simulated daily healthcare utilization series with a known ground-truth intervention effect – a synthetic +12% level shift imposed on March 1, 2025 – to assess how well the estimated counterfactuals recover the true effect. Second, we use counts of weekly injury deaths in Washington State from the US Centers for Disease Control and Prevention (CDC) to study the impact of the June 2021 Pacific Northwest heat dome, treating this extreme weather event as a natural experiment.

`its2s` is maintained on GitHub. All extended documentation can be accessed on the `its2s` website here.

## 4 Case studies

Here we illustrate the utility of `its2s` in two case studies. Case study 1 leverages simulated data to recover the effect of a policy intervention on daily healthcare encounters. Case study 2 uses CDC Wonder data to quantify the impact of the 2021 Pacific Northwest heat dome on injury mortality in Washington state. The case studies are intentionally simple implementations of `its2s`, using predominantly default settings to illustrate the packages’ out-of-the-box utility.

### Casey study 2: simulated policy intervention

#### Background

Case study 1 is a simulation representing a daily time series of healthcare utilization counts for a single geographic unit from January 2022 through December 2026, with a synthetic policy intervention on March 1, 2025. To generate these data, we imposed a baseline of ∼800 visits per day with a moderate upward trend, annual seasonality (peak in early January), day-of-week effects (weekdays higher than weekends), and lower utilization on US federal holidays (∼18% reduction). We sampled daily counts from a Poisson distribution. Beginning on the intervention date, we imposed a constant multiplicative +12% level shift on the rate. We then use `its2s` to estimate the intervention effect,, providing a ground truth against which to evaluate `its2s`.

#### Methods

We use `its2s` with default settings and a Prophet + XGBoost model (default model) to fit the pre-intervention period. We reserved the last 20% of pre-intervention days for a testing set, using all remaining pre-intervention days as training data (default configuration). Our holdout was comprised of all post-event data (2025-03-01 through 2026-12-31). Uncertainty was generated using a MBB (default block length of 14 observations, equal to 14 days on this daily series). We estimated the change in utilization as the difference between the observed utilization count (from simulated time series) and the expected rate (counterfactual predicted from the model we fit on pre-intervention data).

##### Results

During the 671-day post-intervention holdout period (2025-03-01 through 2026-12-31), we estimated an 11.77% excess visit rate (95% eCI: 10.23%, 12.96%), approximately the true 12% level shift. The Prophet - XGBoost model fit the pre-event test time series reasonably well (*MAPE* = 3.9%) (Figure 2), facilitating the recovery of an effect size resembling the true effect.

**Figure 1:**
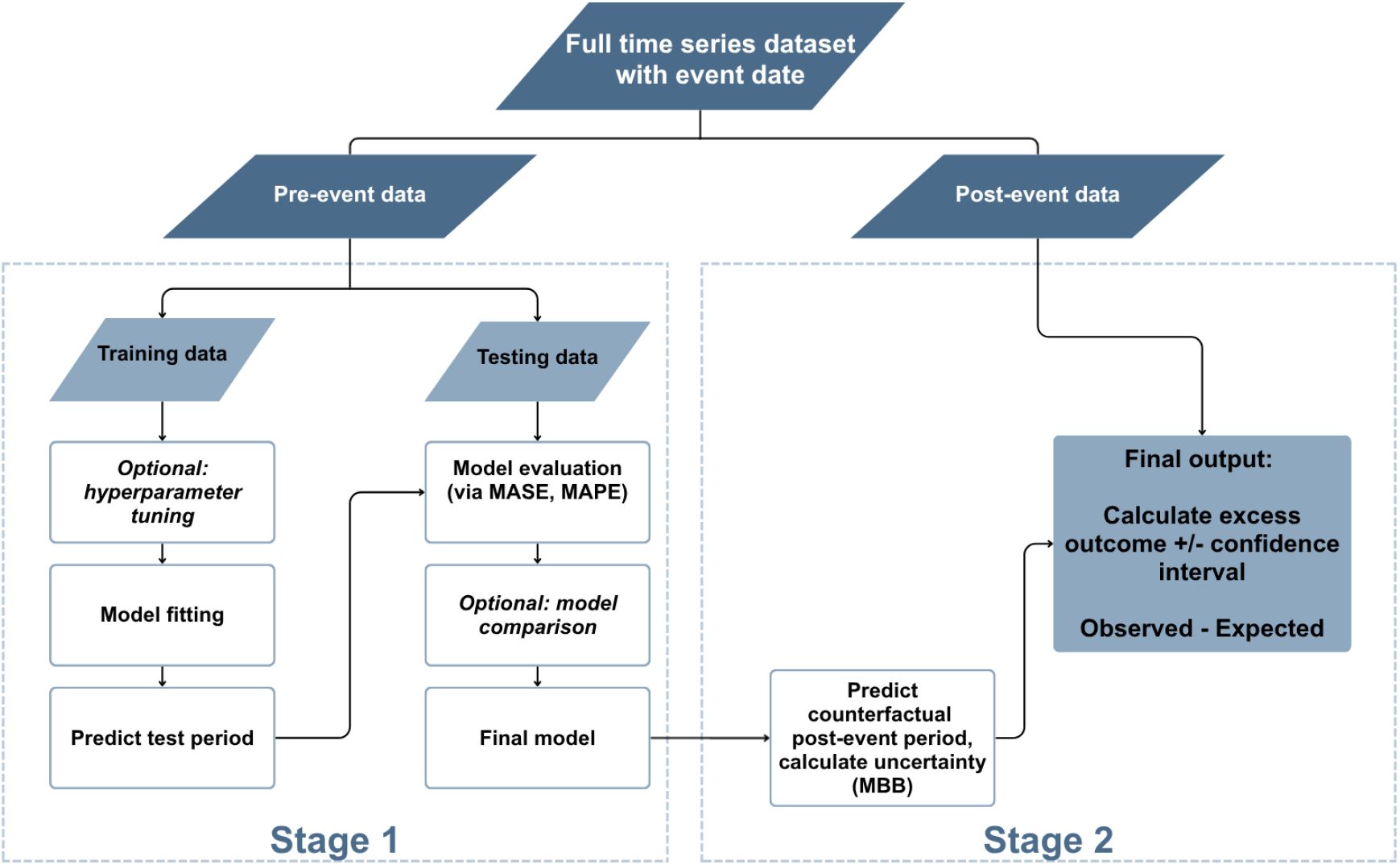
Description of `its2s` analytic workflow. Pre-event data is split into training and testing sets to build the Stage 1 model and predict the counterfactual time series. Stage 2 estimates the difference between the observed post-event time series and the counterfactual time series to estimate the causal effect of the event.

**Figure 2:**
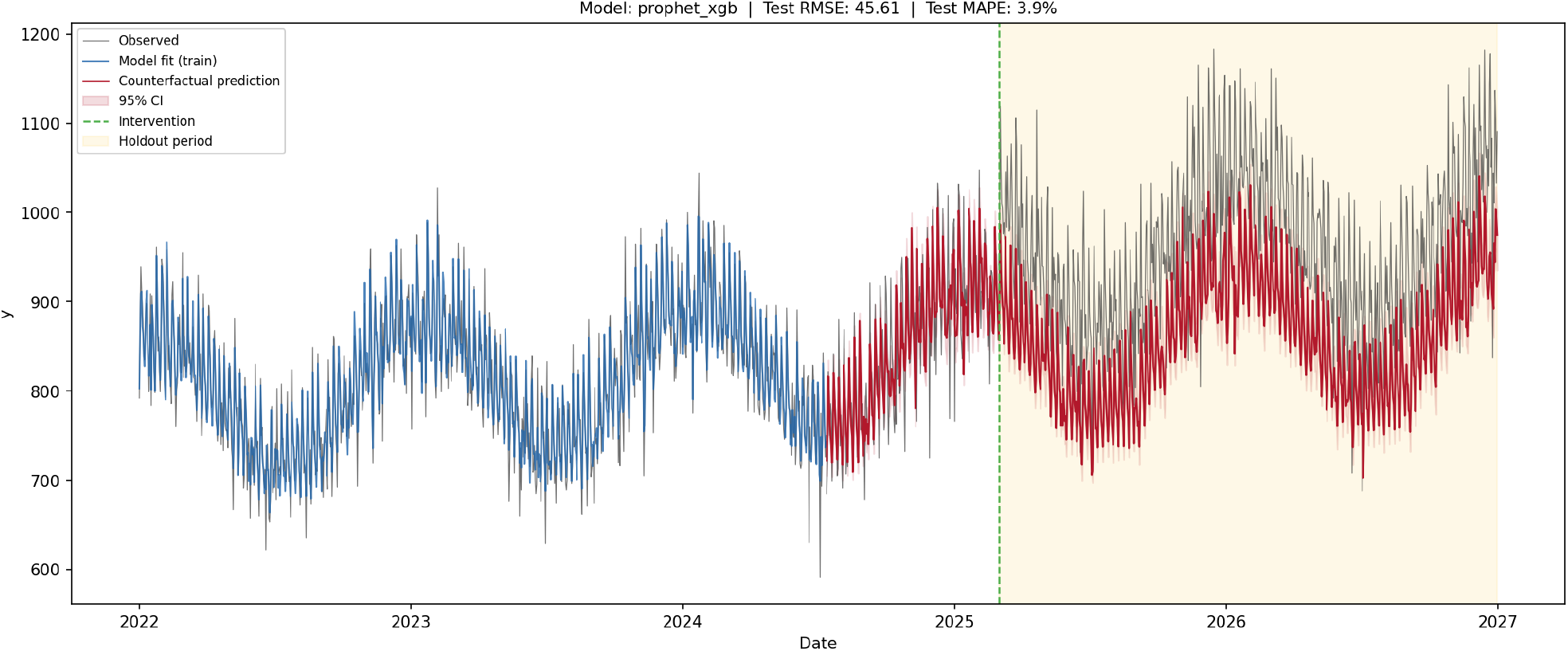
Case study 1: simulated daily data (grey), pre-intervention model fit (blue), and counterfactual prediction with uncertainty for the post-intervention holdout (red; Prophet + XGBoost). Dashed green line represents timing of event.

### Case study 2: 2021 Pacific Northwest (PNW) heat dome

#### Background

In the context of a warming climate, extreme heat is a growing public health concern linked to elevated mortality not only from cardiovascular and natural causes, but increasingly due to injuries (e.g., suicide, drowning, transportation accidents, or occupational accidents in construction or agriculture). ^23–26^ Though historically the Pacific Northwest region of the US has a mild summer, in late June and early July, 2021, the region recorded its highest-ever temperatures due to a ‘heat dome’ where persistent high-pressure atmospheric system trapped and compressed. ^27^ The unprecedented heat among a population with limited previous exposure to such temperatures and relatively little preparedness may have caused substantial excess mortality. ^24,27^ In this case study, we treat the heat dome as a random shock and use `its2s` to apply an ITS analysis to study subsequent changes in weekly injury deaths in Washington State.

#### Methods

We analyzed weekly counts of injury deaths in Washington State from January 2014 through July 2021 from CDC Wonder ^28^ to estimate changes in injury mortality in the 3 weeks following the 2021 Pacific Northwest (PNW) heat dome (intervention date 2021-06-25; holdout weeks beginning 2021-06-26, 2021-07-03, and 2021-07-10). This analysis mirrors that of Casey et al. 2023 (^24^), who used ARIMA modeling. Our case study built a model using most of the default `its2s` settings. Given the strong seasonal temperature trends, we did not use default settings for the train/test split. Instead, we adjusted it to length the training period in order to ensure sufficient training data for the model to capture the seasonality sufficiently. We thus fit a new model on pre-event data (379 pre-event weeks and reserving 11 weeks (78 calendar days, 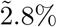 of pre-intervention observations) for out-of-sample testing immediately before 25 June 2021, a deviation from the default). We achieved a reasonable fit on these parameters, though future iterations could adjust the default settings further, too. The counterfactual time series was generated for the 3 weeks following the event, and uncertainty was quantified using MBB. The change in injury deaths was defined as observed minus expected (counterfactual) counts during the post-event holdout period.

#### Results

During the 3-week holdout, we estimated 193 excess injury deaths (95% eCI: 179, 217) relative to the counterfactual, corresponding to a 53% excess (eCI: 49%, 60%). Excess injury-related mortality was highest in the middle week (week beginning 2021-07-03: 104 excess deaths [99, 118]), when observed counts reached 229 versus ∼125 expected. The Prophet-XGBoost model fit the pre-event test time series reasonably well (*MAPE* = 5.5%) (Figure 3).

**Figure 3:**
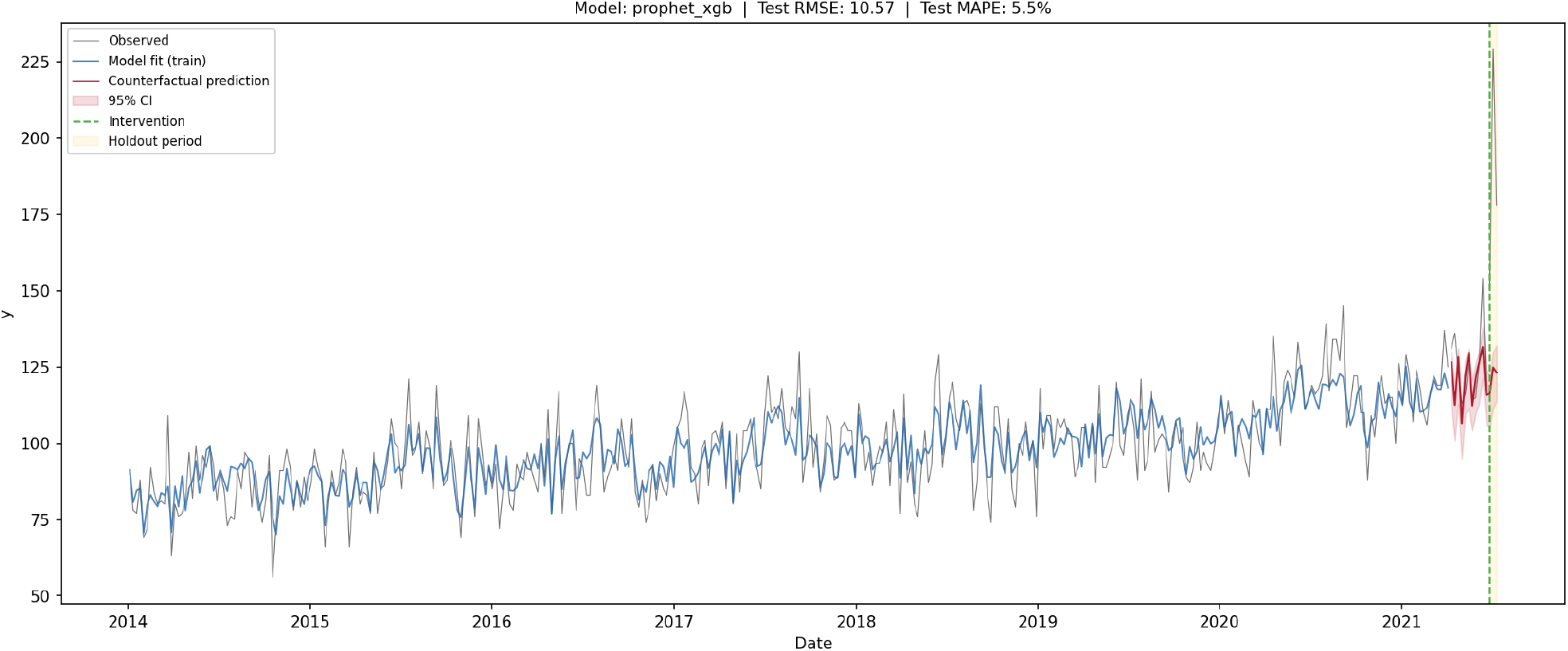
Case study 1: observed weekly injury deaths in Washington State (grey), model fit on pre-intervention data (blue), and counterfactual prediction with uncertainty for the 3-week post–heat-dome holdout (red; Prophet + XGBoost). Dashed green line represents timing of event.

This case study capitalizes on the random timing of the 2021 PNW heat dome to estimate changes in injury mortality attributable to the event. These results are comparable to Casey et al., who reported 159 excess injury deaths (95% CI: 122, 195) over a comparable 3-week window ^24^. Our estimates in this ITS analysis are of similar magnitude and show that `its2s` can be applied in a few lines of code using default workflow settings. This analysis had many limitations – including lack of time-varying covariate adjustment and weekly aggregation, which may mask important daily changes in mortality – but demonstrated a user-friendly implementation of the package for a well-defined shock.

### What we learn from these case studies

Together, these case studies show `its2s` applied with minimal API calls and largely default settings across two complementary settings – a simulated policy intervention in which the package closely recovered a known +12% level shift, and a real extreme-heat mortality application yielding plausible excess estimates. Both examples are illustrative, and are meant to highlight a single workflow for internal validation when ground truth is available. Real-world implementations may range from simple applications as demonstrated here to more complex ones that involve (1) covariate adjustment, (2) custom hyperparameter choices, (3) multi-model comparison frameworks, or (4) dynamic block length selection for estimating uncertainty using MBB.

## 5 Discussion

The objective of employing a two-stage ITS with ML approach is to estimate the effects of an event or intervention by capitalizing on the timing of natural experiments. ITS can be applied to a wide range of natural experiments, including climate shocks (e.g., extreme heat events, wildfires, hurricanes), social or urban policies (e.g., alcohol control policies, protected bike lane) or healthcare policies (e.g., Medicaid expansion). The `its2s` software package allows users to readily implement this study design. The package provides a structured and documented tool for each decision point in the analytical process: `time_series_cv()` for evaluating out-of-sample performance, `compare_models()` for comparing model architectures, `tune_model()` for searching the hyperparameter space along options to use data-driven block-length selection for the bootstrap. `its2s` serves both straightforward use cases, allowing researchers to obtain results quickly with built-in defaults, and advanced use cases with customized pipelines.

Improving model performance – through model selection, covariate specification, or hyperparameter tuning – is the primary lever for strengthening the causal estimates produced by the two-stage ITS approach. ^6,13^ However, achieving good test performance requires researchers to navigate a large and interdependent space of methodological decisions. ^9,11,29^ First, researchers must determine the amount of pre-intervention data to reserve for testing, where too short a test window yields unstable performance metrics, while too long a window leaves too little training data – and the right balance depends on the total length of the pre-intervention series. ^16,18^ Once the data are split, researchers need to choose from different model architectures, each with different assumptions about trend, seasonality, and residual structure. ^6,9,30,31^ Evaluating candidates systematically requires time-series cross-validation, which introduces further structural choices (e.g., the number of folds, the validation window length, and the skip gap between folds needed to prevent information leakage into the held-out test period). ^18,19^ Once a model is selected, searching the hyperparameter space adds another layer as each architecture exposes a distinct configuration space, and evaluating it systematically requires running many cross-validated fits whose cost grows with series length and model complexity. ^9,31^ Further, when estimating uncertainty, for the MBB, the appropriate length should theoretically reflect the autocorrelation structure of model residuals - a structure that varies across series and model specifications. ^9,20,22,32^ This means that a single fixed value for the block length is rarely justified, adding a further analytical decision after the model is fit.

These challenges scale sharply in typical epidemiological applications, where the same analytic steps often need to be repeated across multiple geographic units, age groups or other strata, and outcome types within a single study, and where sensitivity analyses may necessitate varying the training window, cross-validation specifications, or model specifications across an additional set of runs. By automating configuration management, batch processing, and version tracking, `its2s` eliminates much of the time-consuming and error-prone infrastructure burden. This allows researchers to focus on scientific decision making – hyperparameter tuning, model selection, interpretation – rather than maintaining *ad hoc* hard-coded scripts and tracking systems. ^9^

While `its2s` is well-suited for time series analyses with sufficient observations, a clearly defined event or intervention, and data over a relatively long time period (e.g., enough training data to span a full cycle of seasonality), the package has several limitations.

First, this package inherits all the limitations of an ITS design. The validity of the ITS design relies on effective counterfactual estimation, which requires an unverifiable assumption – that pre-event trends would extend into the post-event period absent the event of interest. While strong out-of-sample predictive power is a reasonable indicator of the model’s ability to generalize, it cannot ensure an accurate counterfactual. The design also assumes that no other shock coincides with the event of interest, which only the researcher, not the package, can check. Additionally, the ITS design is not a good fit for studies involving events or interventions with uncertain timing or prolonged duration. From a package perspective, the flexibility of `its2s` risks overfitting – requiring researchers to carefully test and validate models. Additionally, while `its2s` provides functionality to run large batches of different models, it does not handle model-specific customization automatically – this remains a decision for the research team. Finally, while `its2s` provides a computationally efficient framework, computational demands still scale substantially as datasets grow, batches increase, or hyperparameter grids expand.

When exposures cannot be randomized, natural experiments allow researchers to recover causal effects from abrupt exogenous shocks. If the timing of an event is assumed at-random with respect to other outcome determinants, using a population’s own pre-event trajectory can serve as the counterfactual for what would have occurred in the absence of the event – and the difference between the observed and expected outcome trajectories can be interpreted causally. Among the strongest methods for studying these natural experiments is the ITS design. The two-stage ITS with ML approach enhances this method by estimating a counterfactual that flexibly captures complex temporal and seasonal trends. This flexibility introduces methodological and computational barriers to entry that have limited its adoption. We introduce `its2s` as a solution, providing a standardized, reproducible, computationally efficient implementation of the method accessible to all skill levels. `its2s` expands the toolkit for leveraging natural experiments to conduct rigorous, reproducible causal inference – allowing researchers to rapidly and reproducibly quantify the impacts of exogenous shocks and policy interventions.

## Data Availability

All code and data are available in the GitHub repository.

https://github.com/causal-its/its2s

## Acknowledgements

Cursor was used to help draft and refactor portions of the *its2s* codebase. The authors reviewed, tested, and take full responsibility for all code included in the final package.

